# The novel duodenal isolate *Streptococcus salivarius* AGIRA0003 promotes barrier dysfunction and IgG responses in functional dyspepsia

**DOI:** 10.1101/2024.07.15.24310426

**Authors:** Grace L. Burns, Jasmine A. Wark, Emily C. Hoedt, Kyra Minahan, Simonne Sherwin, Jessica K. Bruce, Yenkai Lim, Jing Jie Teh, M. Fairuz B. Jamaluddin, Wai Sinn Soh, Shandelle Caban, Sophie Fowler, Juhura G. Almazi, Ameha S. Woldu, Matthew D. Dun, Pradeep S. Tanwar, Michael D. E. Potter, Erin R. Shanahan, Gerald Holtmann, Mark Morrison, Nicholas J. Talley, Simon Keely

## Abstract

**Background and aims:** Functional dyspepsia (FD) is a highly prevalent disorder of gut-brain interaction (DGBI) that is associated with an altered duodenal microbiota, unexplained low grade duodenal inflammation and altered intestinal permeability. This study aimed to investigate if novel FD-derived bacterial isolates elicited immune responses in FD and the capacity of an immune-stimulating isolate, AGIRA0003 to breach the duodenal epithelial barrier.

**Methods:** Bacterial lysates were investigated for immune reactivity using immunoblotting of patient plasma. Immunoblots were probed with plasma from FD patients (n=44, 46.6±17.5 years, 79.6% female) or controls (n=30, 48.9±15.7 years, 63.3% female). Peripheral gut-homing T cells were quantified by flow cytometry and histological analysis used to investigate duodenal biopsies. Polarised Caco-2 cells and FD duodenal spheroids (n=4 lines) were exposed to *Streptococcus salivarius* AGIRA0003 at a multiplicity of infection of 10 bacterial cells to 1 mammalian cell for 6 hours.

**Results:** The presence of plasma IgG antibodies against *S. salivarius* AGIRA0003 was significantly associated with FD (χ^2^ 15.7, 1, *p*<0.0001). Patients with these IgG antibodies had increased gut-homing lymphocytes (0.33±0.77% vs 1.00±1.46%, *p*=0.046). Strain AGIRA0003, but not related commensal strains, disrupted tight junction proteins in Caco-2 monolayers, and decreased claudin 1 (CLDN1; 0.49±0.11, *p*=0.03), desmocollin 2 (DSC2; 0.64±0.33, *p*=0.03) and desmoglein 2 (DSG2; 0.30±0.12, *p*=0.03) in spheroid monolayers. In addition, DSC2 (2.19±0.97 vs 1.48±0.85, *p*=0.02) and DSG2 (23.22±15.92 vs 12.38±7.34, *p*=0.04) protein levels were decreased in IgG^+^ FD biopsies compared to controls.

**Conclusions:** *S. salivarius* AGIRA0003 is a potential pathobiont capable of impairing duodenal epithelial barrier defences that elicits an immune response in FD patients.

## INTRODUCTION

Functional dyspepsia (FD) is a disorder of gut-brain interaction (DGBI) characterised by recurrent upper gastrointestinal (GI) symptoms with no structural changes identifiable at endoscopy^1^. Given FD is a symptomatic diagnosis, there are no clear biomarkers and diagnosis is often a prolonged process of exclusions. However, FD patients exhibit mucosal “microinflammation”, having greater numbers of duodenal eosinophils^2^, impaired small intestinal barrier integrity and increased mast cell numbers compared to controls^2, 3^. It is thought that microbial and/or dietary antigens interacting with a dysregulated immune system drive FD microinflammation^4, 5^. The role of the duodenum as a major site of antigen sampling^6^ and microinflammation in FD led us to hypothesise that an impaired relationship between the microbiota and immune system would be evident at this site. In that context, FD is associated with compositional changes in the duodenal microbiota, as well as increased bacterial load and altered diversity that can be correlated with meal-related symptoms and reduced quality of life scores^7, 8^. In the FD duodenal mucosa-associated microbiota (d-MAM), *Streptococcus* spp. relative abundance is increased, significantly associated with symptoms^9, 10^ and inversely correlated with *Prevotella* relative abundance^7^. In addition, patients have decreased populations of *Prevotella, Veillonella* and *Actinomyces*^8^. Collectively, associative studies of the d-MAM and FD symptom profile suggest specific microbial species are associated with homeostatic imbalance and disease initiation in FD.

Despite these observations, the nature and consequence of duodenal host-microbiota interactions and their relationship to FD symptoms is not well understood. Decreased barrier integrity, which is a feature of FD^11^, is thought to drive increased translocation of microbes into the mucosa^11^. This might initiate a humoral response to translocating microbes, and thus explain the mucosal microinflammation observed in FD^2, 12, 13^. This hypothesis is supported by our previous work identifying increased intestinal mucosal lymphocytes with a T helper 17-like phenotype in FD patients compared to controls^13^, however, causal antigens for such a response have not yet been identified. As such, this study aimed to investigate immunoglobulins specific to novel d-MAM bacterial species cultured from FD patients. Given *Streptococcus* is the dominant reported genus in the FD d-MAM, associated with more severe symptom burden in other studies^7, 9^, we investigated potential interactions between this genus and the immune system in FD. We demonstrate for the first time that FD patients have plasma IgG responses against *S. salivarius* AGIRA0003. Further, this strain has capacity to alter tight junction and desmosome proteins *ex vivo*, suggesting that *S. salivarius* AGIRA0003 may act as a pathobiont in FD, contributing to immune activation in FD.

## MATERIALS AND METHODS

### Participant selection and recruitment

Participants were recruited from three outpatient gastroenterology clinics (John Hunter, Gosford and Wyong hospitals) in New South Wales, Australia as part of a larger study investigating immune activation in FD^13, 14^. Patients were diagnosed with FD using the Rome III criteria. Outpatient control participants were recruited from those undergoing a screening endoscopy for iron deficiency anaemia (IDA), a positive faecal occult blot test (FOBT), reflux or dysphagia, who exhibited no abnormal pathology. Exclusion criteria and clinical workup is as previously described^13, 14^. During endoscopy, biopsies were collected from the second portion of the duodenum (D2) and approximately 36mL of blood was collected in lithium heparin for the isolation of plasma and peripheral blood mononuclear cells (PBMCs)^13^. Plasma samples from healthy community controls, serum samples from 9 additional outpatient controls and 10 Crohn’s disease (CD) patients with small intestinal inflammation were used from the Digestive Health Biobank (https://digestivehealth.org.au/). All work was carried out with approval from the Hunter New England Local Health District Ethics Committee (references 2019/ETH03893, 2020/ETH03303).

### Culture of bacterial strains

Novel bacterial candidates previously identified through sequencing of the duodenal microbiota in FD compared to controls^8, 10^ were anaerobically isolated from duodenal biopsies, as described^15^. All strains were cultured in heart infusion broth supplemented with hemin (10µg/mL) and vitamin K (0.5µg/mL), before genome sequencing was performed^15^. Cultures were centrifuged and resuspended in sterile PBS at an optical density (OD)_600_ of ∼0.5. Cells were homogenised in Radio ImmunoPrecipitation Assay (RIPA) buffer (Sigma-Aldrich) containing protease and phosphatase inhibitors (HALT cocktail, Thermo Fischer Scientific) to obtained soluble protein lysates.

Probiotic strains *S. salivarius* M18 and K12 (recovered from a commercially available lozenge from Life Extension Florassist®), *S. salivarius* ATCC7073 (glycerol stock provided from Immune Health Research Program, Hunter Medical Research Institute; GenBank: FJ154807b) and *S. salivarius* AGIRA0003 (GenBank: JAHCVC000000000.1) were inoculated in heart infusion medium supplemented with 7.5% mineral solution 2 and 7.5% mineral solution 3^16^, 0.5% yeast extract and 0.1% resazurin (0.1% w/v stock solution) and incubated at 37°C. Growth rates were monitored longitudinally until OD_600_ was 1 for use in cell culture models.

### Immunoblotting for seroreactive immunoglobulin antibodies

Bacterial lysates were electrophoresed (120v, ∼1 hour) using SDS running buffer (1x tris-glycine with 0.1% SDS), on 4-15% polyacrylamide gels (Mini-PROTEAN TGX, BioRad, Hercules, USA). Every second well contained a protein marker. Proteins were then transferred to a polyvinylidene difluoride membrane (90v, 90 mins) and blocked using 2.5% BSA/2.5% skim milk powder (1 hour). Plasma or serum samples (1:500, overnight incubation, 4°C) were used in place of a primary antibody to detect antibodies directed against bacterial proteins, adapted from Lodes *et al*^17^. Blots were incubated for 2 hours with anti-human horseradish peroxidase (HRP) secondary antibodies directed against IgG, IgM, IgA or IgE (Sigma-Aldrich) diluted 1:1000 in blocking buffer. Immunoblots were imaged with a ChemiDoc MP System (Bio-Rad, Hercules, USA) and the presence or absence of banding was recorded for each sample. As there was no way to accurately quantify the specific antibody concentration in the plasma samples, given we did not know the identity of the antigen, absolute quantification of the antibody-antigen relationship was not possible.

### Isolation of seroreactive proteins from total bacterial proteins

Electrophoresis of proteins from bacterial species of interest and a sero-negative control target were conducted as described above. Gel plugs were excised from bands of interest and prepared for liquid chromatography tandem mass spectrometry based on the protocol published by Shevchenko *et al*^18^ **(Supplementary methods)**. The peptide sequences obtained were initially compared against the *S. salivarius* CCHSS3 sequence (due to sequence availability at the time) and the resulting PROKKA-generated protein coding sequences were then manually compared by NCBI BLASTp to the *S. salivarius* AGIRA0003 strain sequence^15^ to identify likely candidates. The complete coding sequence (CDS) for the *S. salivarius* AGIRA0003 “GBS Bsp-repeat domain protein” was then used as a query sequence for a BLASTp-based comparison against all non-redundant GenBank CDS translations + PDB + SwissProt + PIR + PRF (excluding environmental samples from whole genome sequencing projects) database curated by the National Centre for Biotechnology Information (NCBI).

### Caco-2 and duodenal spheroid culture

Polarised Caco-2 monolayers were cultured and maintained as previously described^19, 20^. Briefly, cells were seeded at a density of 1×10^5^ cells/well on Costar Tranwell permeable inserts (pore size 0.4μm polycarbonate membrane, catalogue #3470). Caco-2 monolayers were stimulated in DMEM stimulation media (DMEM high glucose, 1% sodium pyruvate, 1% L-glutamine, 10% FCS) in triplicate for 6 hours at 37°C/5% CO2 with *S. salivarius* strains AGIRA0003, M18, K12 or ATCC7073 at a multiplicity of infection (MOI) of 10, or 1µg/mL LPS. Transepithelial electrical resistance (TEER) was measured at baseline and post-stimulation using an EVOM TEER Measurement device and STX2 handheld chopstick electrodes (World Precision Instruments). Basolateral media was sampled, and spot plated onto heart infusion agar plates to investigate translocation.

FD derived duodenal spheroids^14^ were seeded at a density of 2.5×10^5^ cells/well using in-house L-WRN cell-conditioned media as previously described^21^. Spheroids (n=4 lines) were stimulated in DMEM stimulation media with *S. salivarius* AGIRA0003 multiplicity of infection (MOI) 10 or 1µg/mL LPS for 6 hours (37°C/5% CO_2_) in duplicate. At 6 hours, cells were collected in RIPA buffer, centrifuged and soluble protein fractions collected and stored at −80°C until use.

### Tight junction protein immunoblotting

SDS-Page immunoblotting was used to investigate tight junction protein levels in cell culture samples. Protein samples were normalised to 10µg and electrophoresised at 120v for ∼1 hour before transfer to a PVDF membrane as above. Blots were blocked in 5% BSA for 1 hour, before incubation (overnight 4°C) with the following antibodies in 5% BSA: zonula occludins 1(ZO-1; Invitrogen #61-7300, 1:2000), desmocollin 2 (DSC2; Abcam ab95967, 1: 2500), desmoglein 2 (DSG2; ThermoFisher #PA5-21444, 1:2500), occludin (OCLN; Abclonal A19657, 1:2000), claudin 1 (CLDN1; Invitrogen #51-9000, 1:2000) or β-actin (Abcam #ab8227, 1:10000). Blots were then incubated for 2 hours at room temperature with anti-rabbit IgG HRP secondary antibody (R&D systems HAF008, 1:4000) in 5% BSA and imaged as described above. Protein levels relative to β-actin were determined using ImageJ densitometry analysis and data is presented as fold change to 1.

### Immunohistochemical staining of duodenal biopsies

Sodium citrate buffer (pH 6.0) was used for antigen retrieval in FFPE tissues and following blocking with casein, sections were incubated (overnight 4°C) with anti-ZO1 (Invitrogen #61-7300, 1:200), DSG2 (ThermoFisher #PA5-21444. 1:500) or DSC2 (Abcam ab95967, 1:200). Slides were then incubated with anti-rabbit secondary antibodies (1:500) conjugated to horseradish peroxidase (HRP). Chromogen 3,3’-Diaminobenzidine (DAB) liquid substrate System (Sigma Aldrich, USA) was used to develop sections, which were counterstained with haematoxylin. Slides were digitalised using Aperio AT2 (Leica Biosystems, Wetzlar, Germany) and the DAB staining intensity in each section was scored using the Halo software area quantification algorithm (Indica Labs USA). The pixel intensity score obtained was used to calculate a H-score: H score = (3x % of pixels with strong stain intensity) + (2x % of pixels with moderate stain intensity) + (1x % of pixels with weak stain intensity), allowing for quantitative staining comparison between groups.

### Bioinformatic analysis of AGIRA0003 genome

Identification of unique genes for *S. salivarius* AGIRA0003 was performed with EDGAR v3.0^22^ using NCBI genome deposits for *S. salivarius* M18 (ALIF01000007), *S. salivarius* K12 (AGBV00000000) and *S. salivarius* AGIRA0003 (JAHCVC010000078)^15^. *S. salivarius* AGIRA0003 singleton amino acid sequences were then blasted against the NCBI refseqprotein database attempt annotation of hypothetical genes.

### Statistical analysis

Datasets were analysed and graphed using GraphPad Prism 9 software (GraphPad Software Inc., La Jolla, USA). Data was analysed for normality of distribution using the D’Agostino & Pearson test and parametric/nonparametric testing used depending on outcome. Demographic characteristics were analysed by *t* tests. Fisher’s exact test was used to analyse co-morbidities and confounders. Relationships between the seroreactive response and FD were analysed by Chi-square testing. Immune parameters and paired cell culture data were evaluated by ordinary one-way ANOVA with Holm-Sidak multiple comparison’s test or Kruskal-Wallis test with Dunn’s multiple comparisons testing, t tests with Welch’s correction or Mann-Whitney t test. Figures are presented as mean±SEM, values reported as mean±SD, *p*<0.05 is considered significant.

## RESULTS

### Study cohort

Thirty controls (48.9±15.7 years, 63.3% female) and 44 FD patients (46.6±17.5 years, 79.6% female) were included. Seventeen patients had post-prandial distress (PDS) subtype, 7 met the criteria for epigastric pain syndrome (EPS) and 20 had overlapping EPS and PDS (EPS/PDS). Controls included individuals asymptomatic for FD referred for outpatient endoscopy for symptoms of dysphagia (n=6), unexplained IDA; n=8, unexplained and reflux (n=3) or a positive FOBT (n=3). No evidence of organic GI disease was found during endoscopy or clinical histological examination. In addition, n=10 healthy community were also included. Characteristics of the cohort are presented in **Table 1**. As expected, the proportion of FD patients with co-morbid irritable bowel syndrome (IBS) was significantly higher than the controls (3.3% vs 38.6%, *p*=0.001) and proton pump inhibitor (PPI) use was higher in the FD cohort (10.7% vs 40.6%, *p*=0.02).

**Table 1:** Demographic and clinical characteristics of study cohort.

|  | **Controls** | **FD** | **p value** |
| --- | --- | --- | --- |
|  | n=30 | n=44 |  |
| **Age (mean±SD)** | 48.87 (15.66) | 46.59 (17.46) | 0.57 |
| **Female (%)** | 19 (63.33) | 35 (79.55) | 0.18 |
| **BMI (mean±SD)** | 28.16 (5.80) | 26.93 (5.70) | 0.28 |
| **PPI use (%)#** | 3 (10.71) | 13 (40.63) | 0.02* |
| **H2RA use (%)#** | 0 | 4 (12.50) | 0.12 |
| **NSAIDS use (%)#** | 2 (7.14) | 3 (9.38) | >0.99 |
| **Helicobacter pylori positive (%)#** | 2 (18.18) | 1 (3.57) | 0.187 |
| **IBS co-morbidity (%)#** | 1 (3.33) | 17 (38.64) | 0.001** |

*# denotes a factor that was not provided by all participants included in this cohort*
*BMI = body mass index, PPI = proton pump inhibitor, H2RA = H2 receptor antagonist, NSAIDS = non-steroidal anti-inflammatory drugs, IBS = irritable bowel syndrome*

### *S. salivarius* AGIRA0003 is associated with a humoral response in FD

We selected the most prominent differentially abundant culturable isolates from an FD biopsy and used immunoblotting to screen participant plasma for interactions^15^ (**Figure 1A**). These isolates were taxonomically affiliated with *Streptococcus salivarius* 57.I (hereafter referred to as strain AGIRA0001), *Streptococcus gordonii* Challis CH1 (AGIRA0002) and *S. salivarius* CCHSS3 (strain AGIRA0003). We also investigated an isolate taxonomically affiliated with *Corynebacterium argentoratense* DSM 44202, isolated in parallel with the *Streptococcus* strains. Initial screening of 6 controls and 15 FD patients against these four strains (**Figure 1B**) indicated the presence of bands at ∼75-100kDa and ∼30-35kDa in 3 (50%) controls and 14 (93.3%) FD patients for either AGIRA0001 or AGIRA0003. The presence of any IgG seroreactive banding for either of these *Streptococcus* strains was significantly associated with FD (χ^2^(5.219, 1), *p*=0.02) (**Figure 1C**).

**Figure 1:**
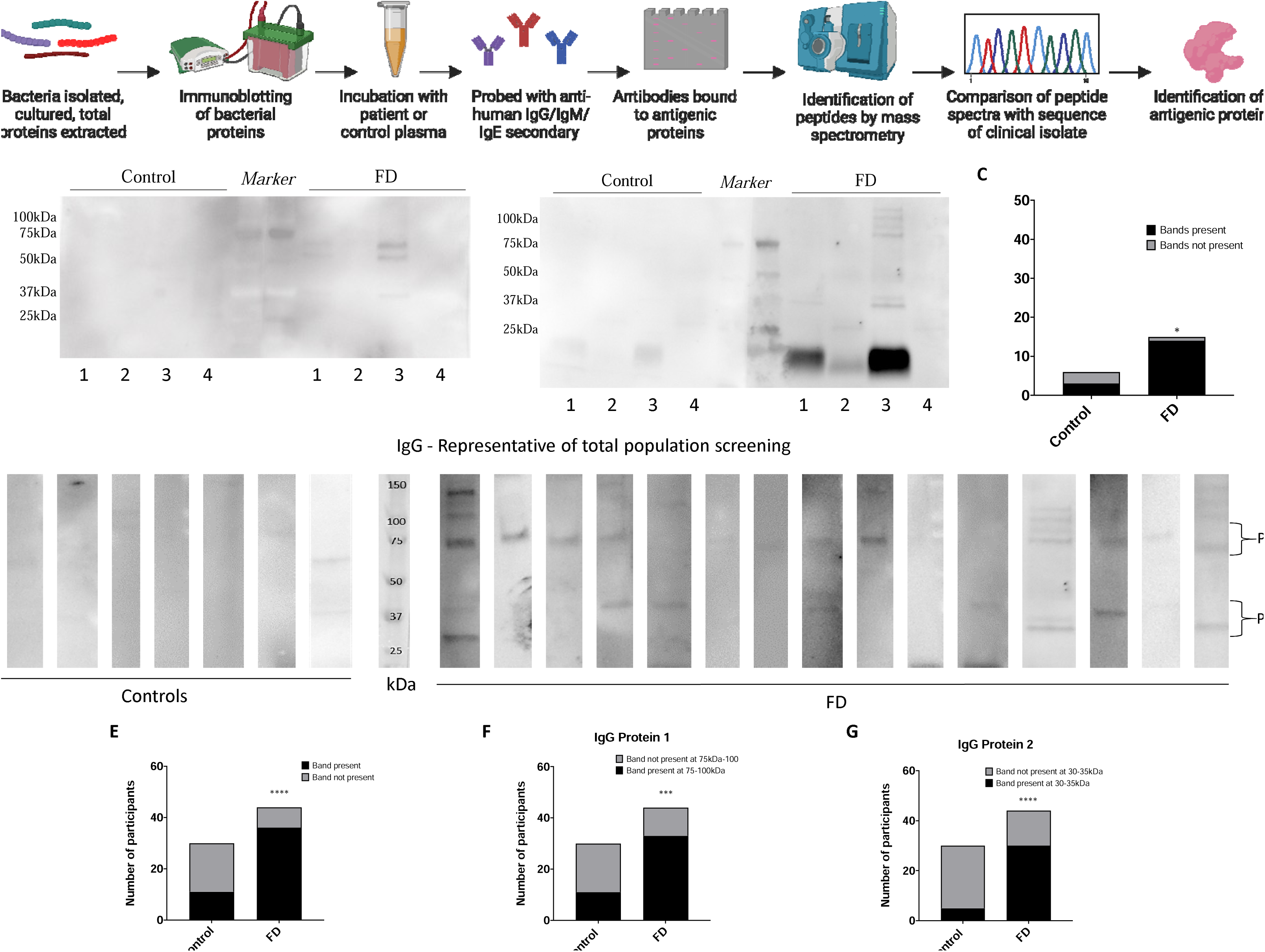
Screening of patient plasma for IgG antibodies against *Streptococcus salivarius* AGIRA0003. Total protein extracted from novel duodenal bacterial lysates was electrophoresed and (A) immunoblotted with patient plasma as the probing antibody to determine interactions between the duodenal microbiota and patient plasma. Figure created with BioRender.com. (B) Representative immunoblots of this screening process, where 1= *Streptococcus salivarius* strain AGIRA0001, 2= Streptococcus gordoni AGIRA0002, 3= *Streptococcus salivarius* strain AGIRA0003, 4**=** Corynebacterium argentoratense. (C) The presence of an interaction with either *Streptococcus salivarius* strain AGIRA0001 or AGIRA0003 was tested for potential associations with FD in the screening population. n=6 controls n=15 FD. The presence of AGIRA0003 IgG seroreactivity in the total cohort was then investigated. (D) Banding patterns observed in immunoblots where total protein extracted from the bacteria was probed with patient or control plasma. (E) The number of FD patients demonstrating a seroreactive response at any molecular weight was compared to seroreactive controls. The number of FD patients with a banding pattern located (F) between 75-100kDa (Protein 1) and (G) between 30-35kDa (Protein 2) compared to the number of positive controls to determine if there was a relationship between this reaction and FD. n=17 controls, n=40 FD patients. Statistical analysis, Chi-square test, \**p*<0.05 \*\*\**p*<0.001 \*\*\*\**p*<0.0001.

Given the visual intensity of banding representing an IgG specific interaction between *S. salivarius* AGIRA0003 and FD plasma upon screening, we focused on this isolate. Representative immunoblots from total cohort are included in **Figure 1D**. In total, 36 (81.8%) FD patients demonstrated a seroreactive response at any molecular weight, compared to 11 (36.7%) controls (χ^2^(15.7, 1), *p*<0.0001) (**Figure 1E**). Within this cohort, 33 (75.0%) FD patients had a banding pattern located between 75-100kDa (hereafter referred to as Protein 1), in contrast to 11 (36.7%) of controls (χ^2^10.9, 1), *p*=0.001) (**Figure 1F**). There was a banding pattern between 30-35kDa (Protein 2) detected in 30 (68.2%) of the FD cohort, compared to 5 (16.7%) of the controls (χ^2^(19.0, 1), *p*<0.0001) (**Figure 1G**). These data validate that there is a significant association between IgG antibodies directed against *S. salivarius* AGIRA0003 and FD. The odds ratio of an IgG response at either molecular weight was 7.8 (95% confidence interval, CI 2.6-20.4), for Protein 1 alone was 5.2 (95% CI 1.9-14.1) and Protein 2 alone was 10.7 (95% CI 3.3-29.7). The sensitivity, specificity, positive and negative likelihood ratios, positive and negative predictive values for an IgG seroreactive response at any molecular weight and each protein individually in FD in included in **Table 2**.

**Table 2:** Sensitivity and specificity for IgG antibodies against *Streptococcus salivarius* AGIRA0003 as a marker of FD.

|  | **IgG seroreactivity**<br>**(95% CI)** | **IgG seroreactivity (Protein 1 only)**<br>**(95% CI)** | **IgG seroreactivity (Protein 2 only)**<br>**(95% CI)** |
| --- | --- | --- | --- |
| Sensitivity | 81.82% (68.04 – 90.49) | 75.00% (60.56 – 85.43) | 68.18% (53.44 – 80.00) |
| Specificity | 63.33% (45.51 – 78.13) | 63.33% (45.51 – 78.13) | 83.33% (66.44 – 92.66) |
| Positive Likelihood Ratio | 2.23 (1.37 - 3.64) | 2.05 (1.24 – 3.37) | 4.09 (1.79 – 9.34) |
| Negative Likelihood Ratio | 0.29 (0.14 - 0.57) | 0.39 (0.22 – 0.70) | 0.38 (0.24 – 0.61) |
| Positive Predictive Value | 76.60% (62.78 – 86.40) | 75.00% (60.56 – 85.43) | 85.71% (70.62 – 93.74) |
| Negative Predictive Value | 70.37% (51.52 – 84.15) | 63.33% (45.51 – 78.13) | 64.10% (48.42 – 77.26) |
| Odds Ratio | 7.77 (2.64 – 20.39) | 5.18 (1.92 – 14.13) | 10.71 (3.34 – 29.70) |

*Wilson/Brown method for 95% confidence interval (CI).*

### The seroreactive response to *S. salivarius* AGIRA0003 is not related to symptom sub-type, co-morbidities or other immunoglobulin responses

To ensure the IgG response against *S. salivarius* AGIRA0003 was not the result of a confounding characteristic, the FD cohort was divided by those who had a detectable IgG response (FD IgG^+^, sero-positive) and those who did not (FD IgG^−^, sero-negative). Correlation analysis was performed for both Protein 1 **(Supplementary Table 1)** and Protein 2 **(Supplementary Table 2)** bands. There was no difference in IgG status for Protein 1 or 2 with regards to *Helicobacter pylori* status, IBS as a co-morbidity, or PPI usage, histamine type 2 receptor agonist (H2RA), or non-steroidal anti-inflammatory drug (NSAIDs) usage. We also screened the serum of 10 Crohn’s disease (CD) patients (40.7±13.8 years, 50.0% female) and 9 additional controls (62.2±13.2 years, 44.4% female). There was no significant relationship between the presence of any IgG response **(Supplementary Figure 1)**, suggesting this phenomenon may be intrinsic to FD. There was no difference in the presence of IgG antibodies against *Streptococcus salivarius* AGIRA0003 between FD patients with a PDS or EPS±PDS subtype, nor did the IgG response prevalence differ between controls recruited from the community compared to the outpatient population. Finally, there was no difference in IgG seroreactivity between FD and FD with concomitant IBS **(Supplementary Figure 2)**.

We also screened a subset of the plasma samples for IgA or IgE antibodies specific to the *S. salivarius* AGIRA0003 strain **(Supplementary Figure 3)**. Probing of 6 controls and 13 FD patients with IgM identified interactions with bacterial proteins between 50-200kDa in 2 (33.33%) controls and 8 (61.54%) FD patients, which were not of statistical significance. There was no significant relationship between IgA seroreactivity against *S. salivarius* AGIRA0003 and FD status and screening for IgE antibodies failed to identify any banding in either group.

### Identification of candidate seroreactive proteins from *S. salivarius* AGIRA0003

Multiple tryptic peptides recovered from the IgG Protein 1 band were matched with the presumptive “GBS Bsp-repeat domain protein” coding sequence predicted from the PROKKA annotation (loci AGIRA0003_00585; NCBI loci equivalent loci MBW4819708.1) of the AGIRA0003 genome (ExPASy bioinformatics resource portal theoretical molecular weight=85.04kDa, theoretical pI=6.49). Additionally, multiple tryptic peptide masses produced from the Protein 2 band were matched with the presumptive “30S ribosomal subunit S2 protein” predicted from the PROKKA annotation (loci AGIRA0003_01557; NCBI equivalent loci MBW4818971.1) of the AGIRA0003 genome (ExPASy bioinformatics resource portal theoretical molecular weight=28.35kDa, theoretical pI=5.04) (**Supplementary Table 3)**.

### Specific d-MAM species are associated with AGIRA0003 seroreactivity in FD

We also investigated if there was any link between d-MAM profile **(Supplementary Methods)** and the presence or absence of IgG antibodies against *S. salivarius* AGIRA0003 in a subsample irrespective of control/FD status (n=20 total, n=4 IgG^-^, n=16 IgG^+^) **(Supplementary Figure 4)**. There were no statistically significant differences in alpha or beta diversity. Spearman’s correlation revealed positive associations with the presence of IgG antibodies against Protein 1 and the relative abundances of *Salmonella*, *Pseudomonas*, *Microbacterium*, *Leifsonia*, *Klebsiella*, *Gemella*, *Fusobacteriaum* and *Denitratisoma* spp. in FD but not control. These data suggest that while overall there is not a specific d-MAM profile generated via 16S rRNA amplicon sequencing associated with *S. salivarius* AGIRA0003 reactivity, there are relationships with specific commensal populations.

### Gut-homing T cells are increased in *S. salivarius* AGIRA0003 reactive FD patients

We investigated if known microinflammatory features we previously reported in a subset of this cohort^12, 13^ were associated with a sero-positive response to the *S. salivarius* AGIRA0003 strain. Duodenal eosinophil counts, gut-homing and duodenal effector lymphocyte profiles were available for 15, 12 and 11 outpatient controls; 36, 33 and 33 FD patients respectively. There was no difference in duodenal eosinophil count (**Figure 2A**) and IgG seroreactive status in controls or FD for Protein 1 (*p*>0.9 for all comparisons) (**Figure 2B**), or for Protein 2 (*p*>0.9 for all comparisons) (**Figure 2C**). IgG^+^ Protein 1 FD patients had a significantly higher proportion of peripheral CD4^+^ gut-homing T cells, compared to controls (0.33±0.77 vs 1.00±1.46, *p*=0.046) (**Figure 2D,E**). For Protein 2, the proportion of gut-homing T cells was approaching significance in IgG^+^ FD patients compared to controls (0.33±0.77 vs 0.89±1.39, *p*=0.057) (**Figure 2F**). There was no difference in duodenal CD4^+^ effector Th2-like or Th17-like cells between seroreactive FD patients or controls **(Supplementary Figure 5)**. These data suggest an association between the presence of a seroreactive response to *S. salivarius* AGIRA0003 and increased proportions of gut-homing T cells in FD patients when compared to controls.

**Figure 2:**
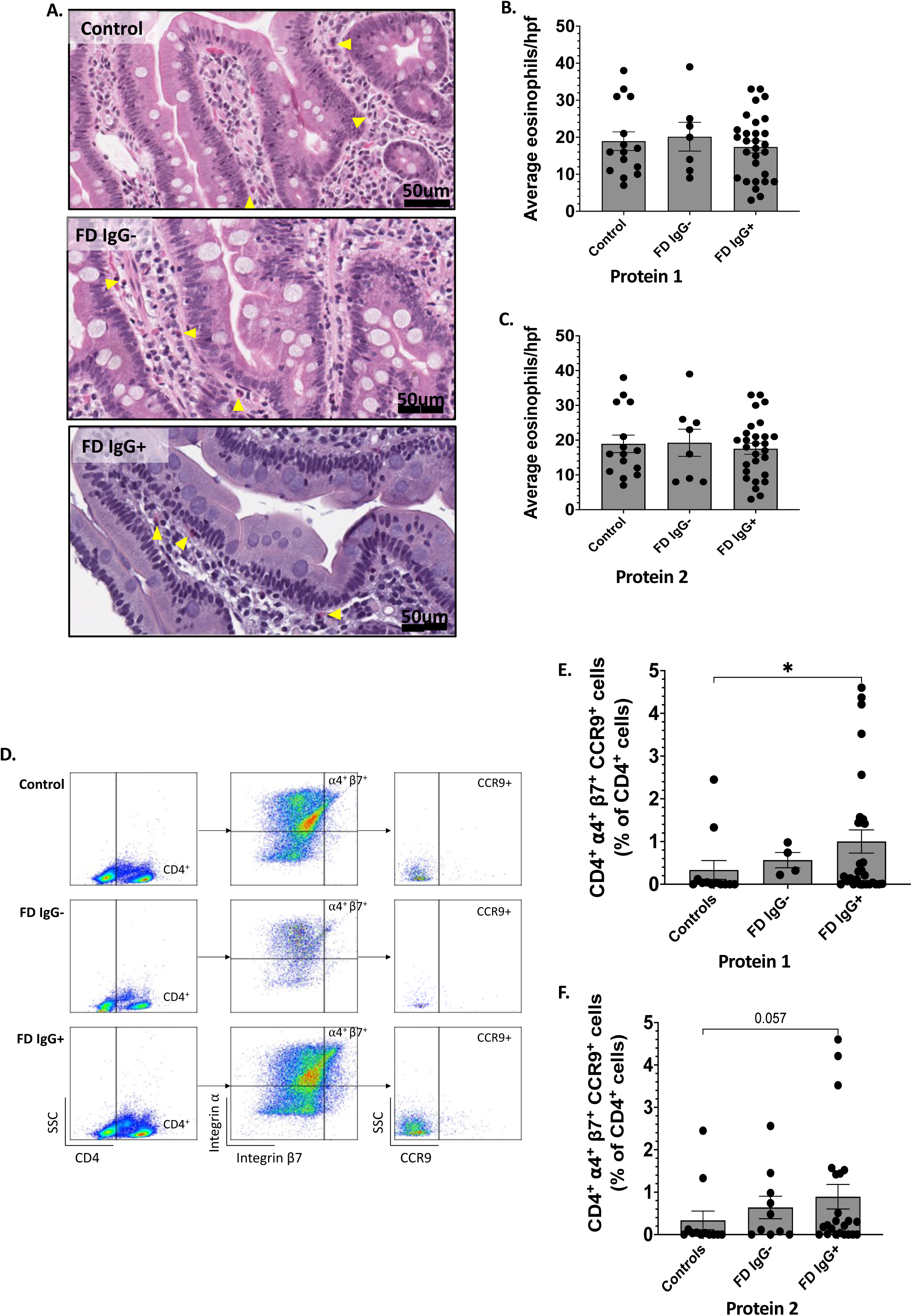
IgG seroreactive status, eosinophils and gut-homing T cells in FD patients. (A) Eosinophils in haematoxylin and eosin-stained biopsies from a subset of the cohort were numerated as part of a previous study. Scale bar = 50um, yellow arrows identify example eosinophils. Eosinophil numbers were compared between controls, IgG^+^ and IgG^-^ FD patients for (B) Protein 1 and (C) Protein 2 (D) Flow cytometry was used to examine gut-homing CD4^+^ cell populations in controls and FD patients in a previous study. These populations were examined in IgG^+^ and IgG^-^ FD patients for (E) Protein 1 and (F) Protein 2. (B,C) n=15 controls, n=36 FD (E,F) n=12 controls, n=33 FD. Data presented as mean±SEM. Statistical analysis: Kruskal-Wallis test, \**p*<0.05.

### *S. salivarius* AGIRA0003 disrupts barrier integrity in Caco-2 monolayers and duodenal spheroids monolayers

To assess the impact of AGIRA0003 on the small intestinal barrier, Caco-2 cell Transwell monolayers were exposed for 6hrs to *S. salivarius* AGIRA0003, as well as related probiotic M18 and K12 strains (**Figure 3A**). Additionally, ATCC7073 was included as a pro-inflammatory control strain isolated from blood. Barrier integrity was assessed using delta transepithelial electrical resistance (ΔTEER) and was significantly decreased in the LPS group (−110.3±12.5 vs −23.99±7.9, *p*=0.004) and AGIRA0003 stimulated cells compared to media (−83.9±27.5, *p*=0.046) (**Figure 3B**). Spot-plating of basolateral media at 3hrs demonstrated greater CFU/mL for AGIRA0003 (*p*=0.02 compared to media) exposed samples in contrast with preparations exposed to the M18, K12 and ATCC7073 strains which were not significant (**Figure 3C**).

**Figure 3:**
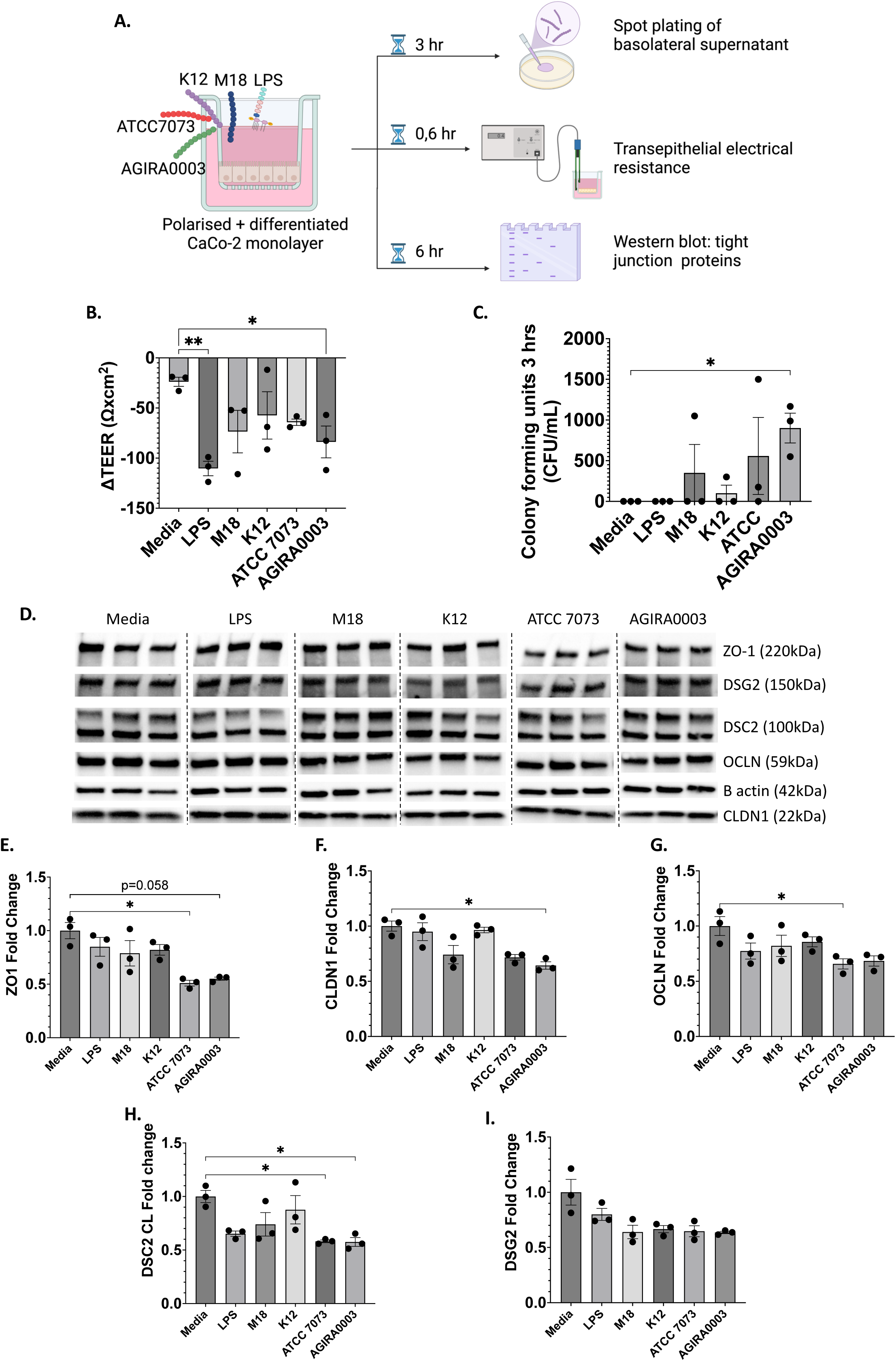
Tight junction proteins following exposure of Caco-2 cells to *S. salivarius* AGIRA0003, M18, K12 and ATCC7073 strains. (A) Caco-2 Transwell monolayers were stimulated for 6hrs with either media only, 1µg/mL LPS, or *S. salivarius* AGIRA0003, M18, K12 and ATCC7073 strains in triplicate. Figure created with BioRender.com. (B) The difference in TEER values (Ωxcm2) of monolayers pre and post 6-hour challenge. (C) Bacterial translocation across the Transwell was assessed by spot-plating from basolateral media at 3 hours post-stimulation and counting resulting colonies. (D) Protein was extracted from Caco-2 cells post-stimulation and tight junction associated proteins (E) ZO-1, (F) CLDN1, (G) OCLN, (H) DSC2 and (I) DSG2 were assessed by immunoblot as fold change to media. Data presented as mean±SEM. Statistical analysis: non-parametric ANOVA with Dunn’s correction. \**p*<0.05, \*\**p*<0.01.

Given tight junction proteins and desmosomes are fundamental to the maintenance of a functional epithelial barrier, we assessed the level of ZO-1, CLDN-1, OCLN, DSC2 and DSG2 (**Figure 3D**). ZO-1 was non-significantly decreased in the *S. salivarius* AGIRA0003 group compared to control (0.55±0.03 AGIRA0003, *p*=0.058) and decreased in the ATCC7073-treated cells (0.51±0.05, *p*=0.01) (**Figure 3E**). CLDN1 was also decreased after exposure to AGIRA0003 compared to control (0.64±0.05, *p*=0.04) (**Figure 3F**), and proinflammatory control strain ATCC7073 decreased OCLN in Caco-2 cells (0.66±0.08, *p*=0.04) (**Figure 3G**). Cleaved DSC2 was decreased in Caco-2 monolayers exposed to ATCC7073 (0.58±0.02, *p*=0.03) and AGIRA0003 (0.58±0.07, *p*=0.04) (**Figure 3H**), while DSG2 level was unchanged across all groups (**Figure 3I**). Collectively, these data suggest AGIRA0003 has potential to compromise the epithelial integrity.

Given tight junction dysfunction is also repeatedly reported in FD, we examined levels of DSC2, DSG2 and ZO-1 in duodenal biopsies from IgG^+^ FD patients. Immunohistochemical staining demonstrated lower levels of mucosal DSC2 (2.19±0.97 vs 1.49±0.85, *p*=0.02) (**Figure 4A**) and DSG2 (23.22±15.92 vs 12.38±7.34, *p*=0.04) in IgG^+^ FD compared to controls (**Figure 4B**). ZO-1 was not significantly changed in IgG^+^ FD compared to control (32.96±19.30 vs 22.77±11.10, *p*=0.08) (**Figure 4C**). A reduction in mucosal DSG2 in FD patients IgG^+^ for Protein 2 was the only difference between IgG^+^ and IgG^-^ FD (21.6±15.1 vs 10.1±6.2, *p*=0.04) for any of the 3 targets **(Supplementary Figure 6)**.

**Figure 4:**
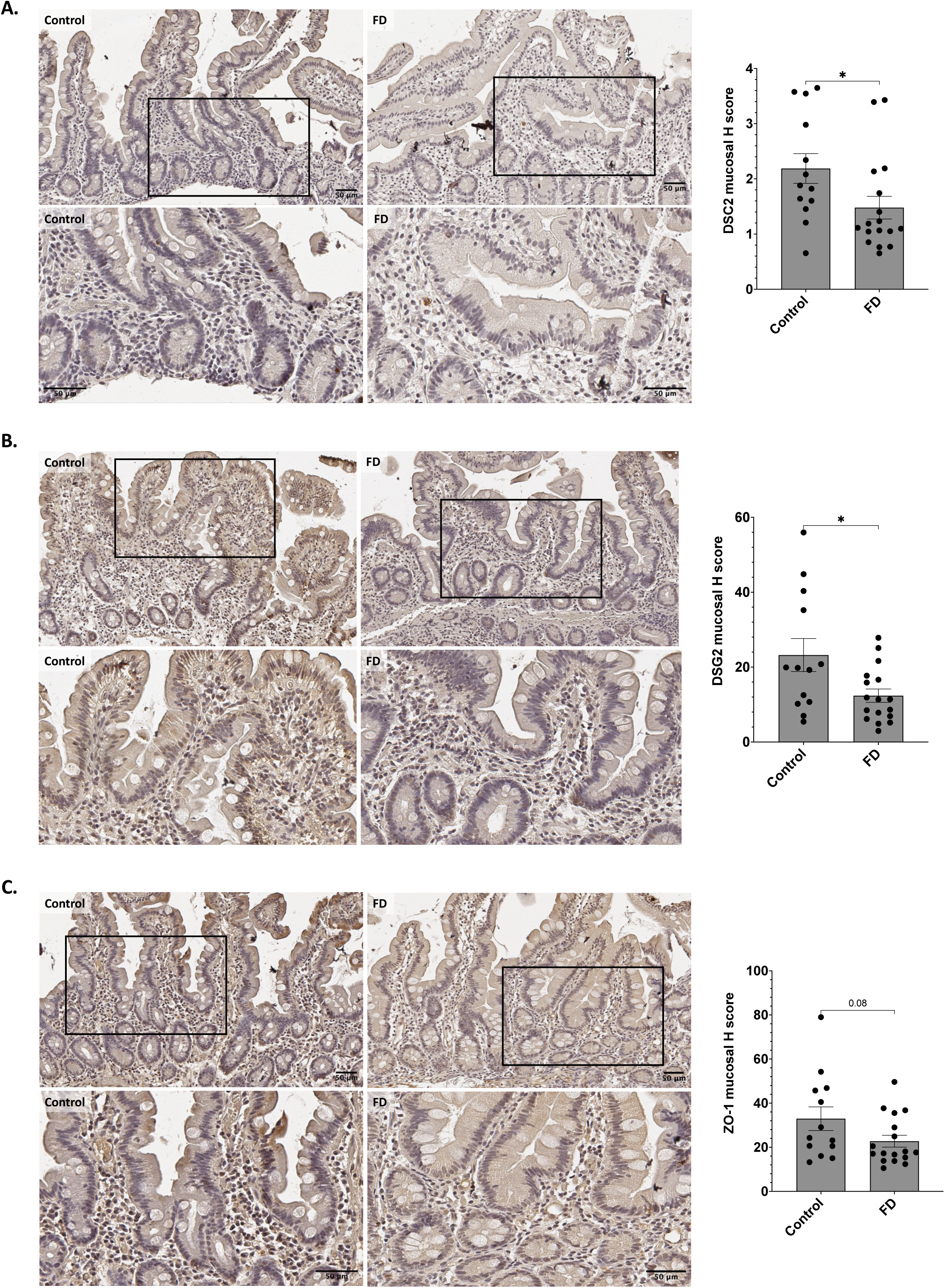
Tight junction associated proteins, DSC2, DSG, and ZO-1, in duodenal biopsies from IgG^+^ FD patients. Formalin fixed, paraffin embedded duodenal biopsies from IgG^+^ FD patients compared to outpatient controls were immunohistochemically stained and H score quantified for (A) DSC2, (B) DSG2 and (C) ZO-1. n=13 controls, n=17 FD. Scale bar = 50µM. Top row=10x magnification, bottom=40x. Statistical analysis: (A) non-parametric t test, (B, C) parametric t test. \**p*<0.05.

We next exposed duodenal spheroids from FD patients with IgG seroreactivity (n=4) to AGIRA0003 for 6 hours and assessed tight junction and desmosomal protein levels by immunoblot (**Figure 5A**). While ZO-1 was unchanged in spheroids exposed to AGIRA0003 (**Figure 5B**), DSG2 was decreased following AGIRA0003 exposure compared to both media (0.30±0.12, *p=*0.03) and LPS (0.99±0.10, *p*=0.03) (**Figure 5C**). CLDN1 was also decreased after AGIRA0003 exposure compared to media alone (0.49±0.11, *p*=0.03) and LPS (0.76±0.23, *p*=0.03) (**Figure 5D**). We also observed a reduction in DSC2 after AGIRA0003 exposure compared to media (0.64±0.33, *p=*0.03) (**Figure 5E**). Collectively, these findings suggest that *S. salivarius* AGIRA0003 actively impairs duodenal barrier defences.

**Figure 5:**
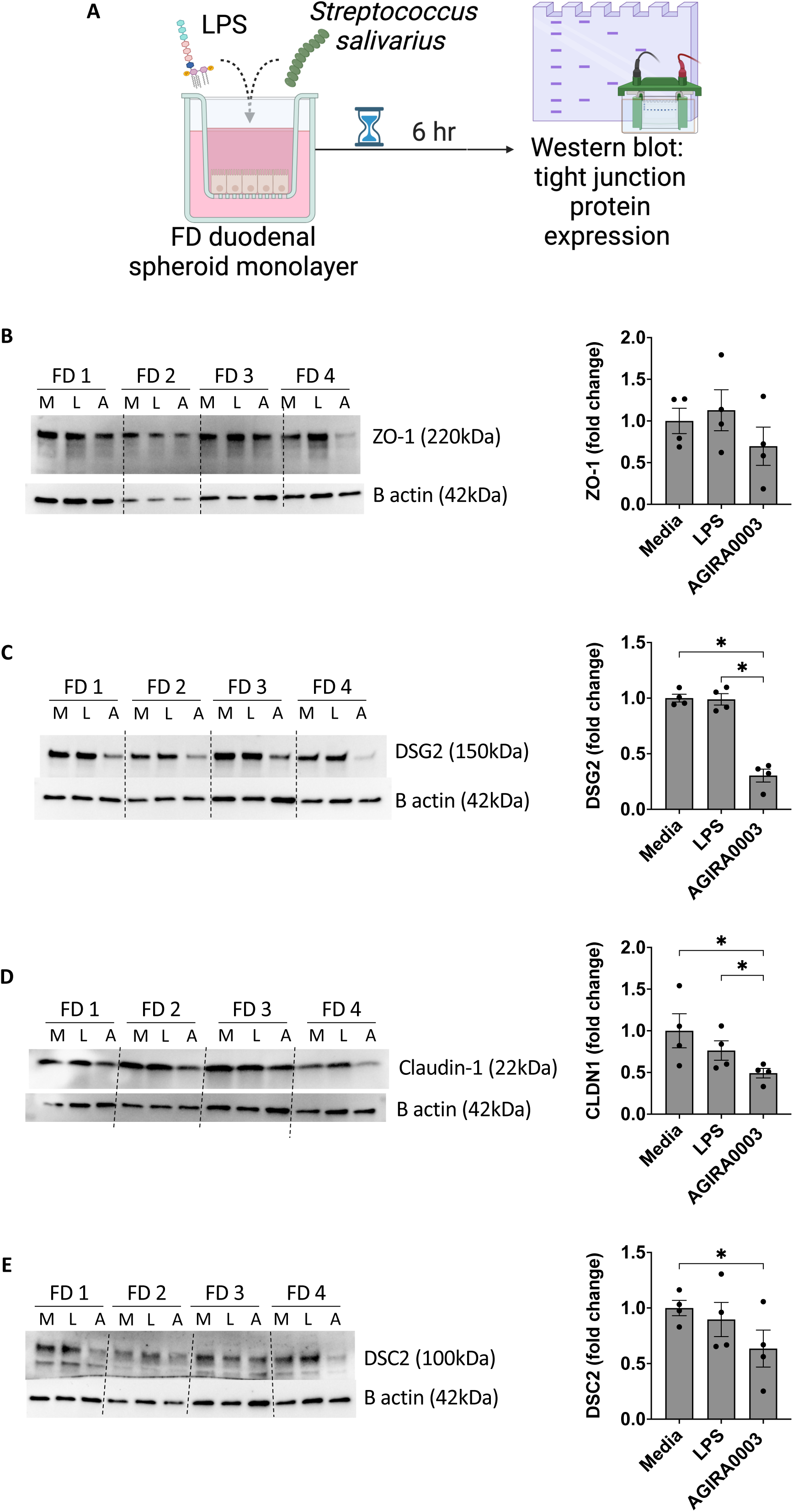
Tight junction proteins following exposure of FD patient-derived duodenal spheroids to *S. salivarius* AGIRA0003. (A) FD patient-derived duodenal spheroids (n=4 lines) were stimulated for 6hrs with either media, 1µg/mL LPS, or *S. salivarius* AGIRA0003 in triplicate. Figure created with BioRender.com. Protein was extracted from the cells at 6 hours post-stimulation and tight junction associated proteins (B) ZO-1, (C) DSG2 (D) CLDN1 and (E) DSC2 were assessed by immunoblot. Data presented as mean±SEM, fold change to 1 (media). Statistical analysis: paired non-parametric ANOVA. \**p*<0.05. M=media only, L=LPS stimulation, A=AGIRA0003 stimulation.

### *S. salivarius* AGIRA0003 may have virulent capacity based on genome interrogation

Given we demonstrated the capacity of *S. salivarius* AGIRA0003 to impact tight junction integrity, we investigated potential pathogenic capacity through *in silico* analysis. Comparative genomics identified 81 unique AGIRA0003 genes when aligned to the probiotic M18 and K12 strains that did not disrupt tight junctions **(Supplementary Table 4)**. Within this list of singletons, 4 genes with virulent properties (e.g., LPS, flagella, pilus, toxins)^23^ were annotated as type II toxin-antitoxin system death-on-curing family toxin, FliM/FliN family flagellar motor switch protein/ type III secretion system (T3SS) cytoplasmic ring protein SctQ [from *Lysobacter enzymogenes*], O-antigen ligase family protein [*Lysobacter* sp. K5869] and sigma 54-interacting transcriptional regulator/type 4 pilus PilR [*Lysobacter enzymogenes*].

## DISCUSSION

In this study, we demonstrate the presence of IgG antibodies in FD patient plasma directed against proteins from *S. salivarius* AGIRA0003, a novel isolate from the FD duodenum^15^, suggesting that *S. salivarius* AGIRA0003 is a pathobiont in the FD mucosal microenvironment. Further, IgG^+^ FD patients had significantly higher circulating gut-homing T cells and these cells have been previously associated with intensity of pain, cramping, nausea and vomiting in FD^24^. Additionally, we show *S. salivarius* AGIRA0003 has capacity to disrupt tight junctions and desmosomes in both polarised Caco-2 cells and FD patient-derived duodenal spheroids. These disruptions are mirrored in the pathology of FD patients, suggesting a potential mechanism for a host-microbe interaction that results in the generation of the observed immune response.

Approximately 10% of FD cases are thought to have a post-infectious aetiology (PI-FD) arising from an acute GI infection^25^. PI-FD is associated with more severe symptoms, including more weight loss, nausea, vomiting and early satiety^26^, and higher incidence of co-morbid insomnia, depression and anxiety^27^ than non-PI-FD. However, it is likely the true prevalence of PI-FD is higher, given the diagnostic reliance on patient recall of infection in the general population. In support of this, anti-cytolethal distending toxin (CtdB) antibodies, produced in response to infection by Gram negative bacteria^28^ are increased in FD and other DGBI patient serum compared to controls^29–31^. DGBI patients also have elevated anti-vinculin antibodies, believed to result from cross reactivity between anti-CtdB antibodies and vinculin proteins^32^, collectively supporting a role for infection in FD onset.

Sequence comparison identified cell wall associated tandem repeats (GBS Bsp-like repeat protein) and a ribosome associated protein (30S ribosomal protein S2) as the likely seroreactive candidates. The ribosomal subunit is important for binding of transfer RNA and messenger RNA during translation^33^. The GBS Bsp-like repeat proteins are implicated in colonisation and interactions of microbes with epithelial cells^34^, including heightened virulence of related *Streptococcus* spp. for competitive advantage in mucosal infections^35^. This GBS Bsp-like repeat protein exists in a tandem repeat sequence in our isolate, in close proximity to a GH25 muramidase catalytic module (AtlA). This is an autolysin of Gram-positive bacteria that hydrolyses 1,4-beta linkages between N-acetylmuramic acid and N-acetyl-D-glucosamine residues to degrade bacterial cell walls^36^. When this protein was overexpressed in Group B *Streptococcus* spp., the cellular morphology was observed to become more elongated and lens-shaped, rather than spherical^37^, suggesting that Bsp-like repeat proteins play a role in mediating cellular morphology, and by extension, evading host detection and enabling mucosal colonisation^35, 38^.

Tight junction proteins are integral in forming the intestinal epithelial barrier and inhibiting the translocation of mucosal antigens through the paracellular space to the lamina propria. Given that our cell culture data demonstrates the capacity of *S. salivarius* AGIRA0003, but not the related probiotic strains we assessed, to translocate through intestinal monolayers while reducing tight junction and desmosome protein levels in both Caco-2 and FD-derived spheroids, this may represent a pathway for immune sensitisation and activation in FD. Many entero-pathogens, have adapted to target or disorganise tight junction proteins to allow for epithelial translocation via virulence proteins^39^. Comparative genomics analysis of the *S. salivarius* AGIRA0003 genome identified 4 unique genes with potential virulence. Of particular interest was the FliM/FliN family flagellar motor switch protein/Type 3 secretor system (T3SS) cytoplasmic ring protein SctQ. The T3SS is a transmembrane complex that enables protein transport into host cells and is a feature of highly pathogenic bacteria including *Enterococcus, Shigella, Vibrio, Salmonella* and *Pseudomonas*^40^. This complex is integral to the capacity of such enteropathogens in targeting tight junction proteins^41, 42^. The Walkerton, Ontario, Canada outbreak of acute gastroenteritis, due to T3SS complex and/or FliM/FliN proteins^43, 44^ positive *Escherichia coli* 0157:H7 and *Campylobacter* spp.^45–47^, resulted in a higher prevalence of PI-FD up to 8-years post outbreak in those who contracted gastroenteritis^45^.While the complete secretion system was not encoded in the *S. salivarius* AGIRA0003 genome, if functionally expressed, this protein may act as an antigenic stimulant of the host response in patients with previous history of infection with other bacterium utilising this system, potentially contributing to the onset of PI-FD.

Desmosomes (including DSC2, DSG2) are junctions critical for strong adhesion and linking cytoskeletal filaments to cell-ceil contact sites, and as such, disruption to these proteins weakens the epithelial monolayer, a feature leveraged by some pathogens with capacity to disassemble desmosomes^48^. For example, proteolytic cleavage of desmoglein proteins by *S. pyogenes* has been associated with impaired epidermal barrier and the development of cutaneous infection^49^. Further, DSC2 is targeted by *Giardia duodenalis*^50^ and loss of DSG2 has been implicated in Crohn’s disease^51^. Reduction in DSC2 and DSG2 gene expression^52^, and DSG2 protein levels^11^ have been shown in duodenal biopsies from FD patients compared to controls. As such, our findings proposition *S. salivarius* AGIRA0003 as a FD pathobiont capable of disassembling desmosomes and tight junction proteins. This may represent a microbial driven mechanism for the reduced barrier integrity in FD patients that would then permit increased interaction of luminal contents with the host immune system.

While we have shown that *S. salivarius* AGIRA0003 drives barrier dysfunction and that IgG seroreactivity is associated with gut homing T cells, a larger study (incorporating a known and unselected validation cohort) across multiple international sites is required to conclusively establish *S. salivarius* AGIRA0003 as a causative agent and biomarker for FD. Nevertheless, our cohort was recruited from three sites across New South Wales, Australia spanning well over a 100km radius. Further, we have shown that AGIRA003 seroreactivity is not associated with Crohn’s disease, where patients exhibit barrier dysfunction nor outpatient controls. While biomarkers including zonulin, anti-CtdB, and anti-vinculin antibodies have been proposed to distinguish DGBIs^29, 30, 53, 54^, their translation into standard care is impeded by heterogeneity and marker overlap with other diseases/physiological states^55^. Thus, a larger validation cohort may provide utility for *S. salivarius* AGIRA0003 as a diagnostic marker discriminating FD from other conditions (including inflammatory bowel diseases and coeliac disease). A further limitation is the lack of data on the absolute abundance of *S. salivarius* AGIRA0003 globally, as most data assessing small intestinal mucosal sites has utilised 16S RNA gene amplicon sequencing, which does not allow for strain level identification^56^ and cost and difficulty removing host DNA from duodenal samples limits shotgun metagenomics. To that end, progress has been made with capturing and characterising the MAM by “culturomics’ a novel combination of microbe culture with metagenomic sequencing^57^. Here, our findings show that microbe cultivation also represents a meaningful and feasible alternative to functionally examine the d-MAM, beyond the widely practised “culture-independent” methods. Investigation of the prevalence of this strain in other intestinal niches, in addition to the duodenum, will facilitate further understanding of its role in FD pathophysiology.

We have provided evidence of a dysregulated relationship between the novel duodenal *S. salivarius* strain AGIRA0003 and the host immune system in FD. Relationships between GI disease and immune responses involving the commensal microbiota have been previously reported, although to our knowledge, ours is the first study to identify seroreactivity to a specific bacterium in the blood of FD patients. Our comparative genomic analysis of other *S. salivarius* strains demonstrated that *S. salivarius* AGIRA0003 contains unique genes putatively implicated with virulence and we have functionally highlighted the capacity of this strain to reduce barrier integrity via disruption of tight junction and desmosome proteins. Importantly, our data show that duodenal pathobionts can cause the epithelial dysfunction previously identified in FD^11^ and that immunoreactivity to pathobionts can explain the immunological features of FD^13, 24^. Overall, our findings suggest *S. salivarius* AGIRA0003 is a novel pathobiont that may contribute to FD pathogenesis, and thereby represent a potential biomarker and therapeutic target.

## Data Availability

: All data are available in the main text or the supplementary materials, further information is available from the authors upon reasonable request. S. salivarius AGIRA0003 sequence details can be found in E. C. Hoedt, E. R. Shanahan, S. Keely, A. Shah, G. L. Burns, G. J. Holtmann, N. J. Talley, M. Morrison, Draft Genome Sequence of Streptococcus salivarius AGIRA0003, Isolated from Functional Gastrointestinal Disorder Duodenal Tissue. Microbiol Resour Announc 10, e0075821 (2021).

## Acknowledgments

We gratefully acknowledge the significant contributions of the late Prof. Marjorie M. Walker to the conceptualization, supervision, reviewing, and editing of this manuscript, as well as her profound impact on the fields of DGBI and duodenal pathologies.

We thank the Hunter Medical Research Institute Core Histology Facility for processing, sectioning, and scanning histological sections. The authors wish to thank the Department of Gastroenterology at the John Hunter Hospital, New Lambton Heights, Australia and Newcastle Endoscopy Centre, Charlestown, Australia for their support and assistance with the collection of biopsy samples.

The authors generously thank Dr. Kurtis Budden (Immune Health Research Program, Hunter Medical Research Institute and University of Newcastle) for providing a stock of *S. salivarius* ATCC7073.

## Author contributions

Conceptualization: GLB, MM, GH, MMW, NJT, SK

Methodology: GLB, JAW, ECH, MM, NJT, SK

Investigation: GLB, JAW, ECH, KM, SS, JKB, SF, YL, JJT, MFBJ, WSS, SC, JGA, ASW, MDP, ES

Visualization: GLB, JAW

Resources: MDD, PST, GH, MM, NJT, SK

Writing – original draft: GLB, JAW, SK

Writing – review & editing: All authors

## Data and materials availability

All data are available in the main text or the supplementary materials, further information is available from the authors upon reasonable request. *S. salivarius* AGIRA0003 sequence details can be found in E. C. Hoedt, E. R. Shanahan, S. Keely, A. Shah, G. L. Burns, G. J. Holtmann, N. J. Talley, M. Morrison, Draft Genome Sequence of Streptococcus salivarius AGIRA0003, Isolated from Functional Gastrointestinal Disorder Duodenal Tissue. Microbiol Resour Announc 10, e0075821 (2021).

