## Supplementary methods and data for "The novel duodenal isolate *Streptococcus salivarius* AGIRA0003 promotes barrier dysfunction and IgG responses in functional dyspepsia"

**Burns, Wark et al Supplementary Materials**

**Supplementary methods**

Isolation of seroreactive proteins from total bacterial proteins

Two identical gels were run in tandem per bacterial target, to allow one to undergo fixation and overnight staining with Sypro Ruby; while the other underwent immunoblotting, as described above with FD patient $IgG^+$ plasma. The immunoblot image identifying specific bands of interest was overlayed on the total protein image taken from the corresponding Sypro Ruby stained gel. Gel plugs from the corresponding area in the protein gel were excised using a scalpel blade and were also collected from the sero-negative bacterial lysate gel. Protein free areas of both gels were obtained for background subtraction during mass spectrometry. Gel plugs were de-stained, washed and dehydrated using 100% acetonitrile vacuum centrifugation and prepared for mass spectrometry. Briefly, sequencing grade trypsin (Promega, Madison, Wisconsin, USA) and ammonium bicarbonate solution was used for the re-hydration and overnight peptide digestion of gel plugs. 10% trifluoroacetic acid was used to quench trypsin, and sample supernatants were collected and stored at -20°C following sonication prior to analysis.

Liquid chromatography tandem mass spectrometry (LC-MS/MS)

Mass spectrometry was performed using a Q-Exactive Plus hybrid quadrupole-Orbitrap MS system (Thermo Fisher Scientific, Bremen, DE) coupled to a Dionex Ultimate 3000RSLC nanoflow HPLC system. Samples were loaded onto an Acclaim PepMap100 C18 75µm x 20mm trap column (Thermo Fisher Scientific) for pre-concentration and online desalting. Separation was then achieved over an EASY-Spray PepMap C18 75µmx250mm column (Thermo Fisher Scientific, Bremen, DE), employing a linear gradient from 2% to 32% acetonitrile at 300nl/min over 120 min. Data dependent acquisition was performed on the Q-

Exactive Plus MS System operated in full MS/data-dependent MS/MS mode. A precursor ion of endogenous peptide was measured in the Orbitrap scanning the mass range from m/z 390-1400 with an Orbitrap resolution of 70,000, a target automatic gain control value of 1e6, and maximum fill times of 50ms. The 20 most intense multiply charged precursors were selected for higher-energy collision dissociation (HCD) fragmentation with a normalised collisional energy (NCE) of 30, the MS/MS fragments were measured at an Orbitrap resolution of 17,500 AGC of 5e5, and maximum fill times of 110ms.

Gut-homing T cells and eosinophil analysis

$CD4^+$ and $CD8^+$ gut-homing T cells in PBMCS, duodenal eosinophil counts in haematoxylin and eosin-stained in formalin fixed, paraffin embedded (FFPE) sections and duodenal $CD4^+$ effector lymphocyte populations were previously reported[1]. Where matched samples were available, we re-analysed this data to investigate these cell populations in FD patients with and without $IgG^+$ responses to *S. salivarius* AGIRA0003.

16S rRNA amplicon gene sequencing

Duodenal biopsies (1 per participant) collected in RNAlater (Thermofisher Scientific) were first lysed through bead homogenization using the Precellys24. The total DNA was then extracted and purified with the Promega Maxwell automated DNA recovery system following manufacturer's instructions. The 16S rRNA gene encompassing the V6 and V8 regions was targeted using the 917F (5'-GAATTGRCGGGGRCC;-3') and 1392wR (5'-ACGGGCGGTGWGTRC;-3') primers modified to contain Illumina specific adapter sequence. The 16S library was constructed following the Illumina protocol #15044223 Rev.B (Illumina, Inc., United States) by the Australian Centre for Ecogenomics (ACE). Indexed amplicons were pooled together in equimolar concentrations and sequenced on MiSeq

Sequencing System (Illumina) using paired end sequencing with V3 300bp chemistry. Passing quality control (QC) of resulting sequence was determined as 10,000 raw reads per sample prior to data processing and passing QC metrics in line with Illumina supplied reagent metrics of overall Q30 for 600bp reads of >70%.

Microbiota analysis

A total of 20 samples were sequenced under one sequence run. Data was processed with QIIME2 (v2020.11) and DADA217. Taxonomic assignment was performed in R (v4.1.3) with DADA2 (v1.20.0) against SILVA SSU r138 reference database18. Analysis of microbiota diversity was performed in R, using packages phyloseq (v1.38.019), breakaway (v4.7.3), ampvis2 (v2.7.620), and microbiome (v1.16.0). Rare taxa that had less than 0.01 percent relative abundance across all samples were removed. Alpha diversity was analysed for significant difference through Wilcoxon test, Bonferroni correction was used for post-hoc comparisons. Adonis2 (PERMANOVA) was used to test statistical significance of Bray-Curtis PCoA. Data was normalised with total sum scaling and subsequently transformed using centre log ratio for rank sum testing.

**Supplementary Table 1: Potential correlations between seroreactivity to *Streptococcus salivarius* AGIRA0003 75-100kDa and demographic information**

| | FD IgG -ve for Protein 1 | FD IgG +ve for Protein 1 | *p* value |
|---|---|---|---|
| *Helicobacter pylori* infection (%) | 0/7 (0.00) | 1/21 (4.55) | >0.99 |
| IBS co-morbidity (%) | 5/11 (45.45) | 12/27 (44.44) | >0.99 |
| PPI usage (%) | 4/9 (44.44) | 10/23 (43.48) | >0.99 |
| H2RA usage (%) | 1/9 (11.11) | 3/23 (13.04) | >0.99 |
| NSAIDs usage (%) | 1/9 (11.11) | 2/23 (8.70) | >0.99 |

*Fisher's exact test. Data not available for all included samples, /n for each category. IBS = irritable bowel syndrome, PPI = proton pump inhibitor, H2RA = histamine type 2 receptor antagonist, NSAIDs = non-steroidal anti-inflammatory drugs*

**Supplementary Table 2: Potential correlations between seroreactivity to *Streptococcus salivarius* AGIRA0003 30-35kDa and demographic information**

| | **FD IgG -ve for Protein 2** | **FD IgG +ve for Protein 2** | ***p* value** |
|---|---|---|---|
| *Helicobacter pylori* infection (%) | 0/7 (0.00) | 1/21 (4.76) | >0.99 |
| IBS co-morbidity (%) | 6/13 (46.15) | 11/25 (44.00) | >0.99 |
| PPI usage (%) | 2/10 (20.00) | 12/22 (54.55) | 0.12 |
| H2RA usage (%) | 1/10 (10.00) | 3/22 (13.64) | >0.99 |
| NSAIDs usage (%) | 0/10 (0.00) | 3/22 (13.64) | 0.53 |

*Fisher's exact test. Data not available for all included samples, /n for each category. IBS = irritable bowel syndrome, PPI = proton pump inhibitor, H2RA = histamine type 2 receptor antagonist, NSAIDs = non-steroidal anti-inflammatory drugs*

**Supplementary Table 3: Protein sequences for identified seroreactive proteins in *Streptococcus salivarius* AGIRA0003 clinical isolates**

| Protein | Sequence |
|---|---|
| PROKKA_00585 GBS Bsp-like repeat protein | MRTKDFIYYASAAVLLAVTTQVAQADEVATTKTPSVTEENQYQSATAAEIFGGEAALPVTPKSTVSAPAATSE<br>VAKASAPAVSMSPASQSSEAATASTSVTSSVVSSESATASTSATSSETSNSAVATPAKLTNSTDVPSQTLKVQP<br>K**TFIDVSSHNGDISVDDYRALAR**QGVGGVVVKLTEDTWYNNPKAPSQVR**NAQIAGLQVSTYHFSR**YTTEE<br>EARAEAR**FYIQAAQKLNLPK**STVMVNDFEDSNMLPNINR**NTQAWVNEMRK**HGYNNLMFYTSASWLDENN<br>LGYRGPVSTSQFGIENFWVAQYPSSSLTATSAKNMRYNAKTGAWQFSATANLLPGKHVFDQSVDYTGRFTA<br>NASVEADPTQGDLSGTISIVNNNPTLGSFDVVISNVKAPNGVQTVSVPIWSEINGQDDIIWYTANRQNNGTYTV<br>NVKASAHKNSTGLYNVHLYYVQKDGQLTGVGGTTTQVFIGKKPEISVFANLSISKNENNGTFTIIAKNLK**GLE**<br>**GYKEVK**IPFWSHANGMSDIK**WYTPTRQADGSYTVTVK**ASDHENANGRYEAQVFYIDARGQKRFVQKAFV<br>ERNDPSKPTGVISITNNKDSGTFDVVISDVYSPKGVRTVQVPIWSEVDGQDDIRWYEATRQTDGNYKVTVQV<br>ANHKYSTGIYNVHLYYIQNDGSQIGVGGTQTNVTLSEPKADLAITGLNNATGSYDVVISNLVAPRGFKEVLVP<br>TWSEKNGQDDIIWYKAAK**QANGDYKVTVR**SSNHKGDSGLYNSHVYLVDNDGKFIGLGGKSVTLKRA |
| PROKKA_01557 30S ribosomal protein S2 | MAVISMKQLLEAGVHFGHQTRRWNPKMAK**YIFTERNGIHVIDLQQTVKMADTAYEFVREAAANDAVILF**<br>**VGTKKQAAEAVAEEATRAGQYYINHR**WLGGTLTNWDTIQKRIARLKEIKQMEADGTFEVLPK**KEVALLN**<br>**KQR**ARLEK**FLGGIEDMPR**IPDVIYIVDPHKEQIAVKEAKKLGIPVVAMVDTNADPDEIDVIIPANDDAIRAVK<br>LITSKMADAIIEGNQGEDASADFQEAAAADSIEEIVEVVEGDNN |

***Bold** represents tryptic peptide masses obtain from mass spectrometry data and where they match the coding sequence in the S. salivarius AGIRA0003 open reading frames.*
*Underlined amino acid sequences represent distinct peptides identified by mass spectrometry analysis that are derived from the larger tryptic fragment*

### Supplementary Table 4: Singleton genes within the *Streptococcus salivarius* AGIRA0003 genome compared to *S. salivarius* M18 and K12 strains.

| Singleton: | Protein: | Description | Scientific Name | Max Score | Total Score | Query Cover | E value | Per. Ident | Acc. Len | Accession |
|---|---|---|---|---|---|---|---|---|---|---|
| KJE23_00305 | site-specific integrase | | | | | | | | | |
| KJE23_00315 | hypothetical protein | | | | | | | | | |
| KJE23_00320 | replication initiation factor domain-containing protein | | | | | | | | | |
| KJE23_00325 | hypothetical protein | | | | | | | | | |
| KJE23_00330 | helix-turn-helix domain-containing protein | | | | | | | | | |
| KJE23_00335 | tetratricopeptide repeat protein | | | | | | | | | |
| KJE23_00340 | hypothetical protein | | | | | | | | | |
| KJE23_00345 | hypothetical protein | | | | | | | | | |
| KJE23_00350 | hypothetical protein | | | | | | | | | |
| KJE23_00575 | hypothetical protein | | | | | | | | | |
| KJE23_00770 | peptide ABC transporter substrate-binding protein | | | | | | | | | |
| KJE23_00775 | ABC transporter permease | | | | | | | | | |
| KJE23_02000 | hypothetical protein | | | | | | | | | |
| KJE23_02235 | hypothetical protein | | | | | | | | | |
| KJE23_02325 | type II toxin-antitoxin system Phd/YefM family antitoxin | | | | | | | | | |
| **KJE23_02330** | **type II toxin-antitoxin system death-on-curing family toxin** | **Death on curing protein, Doc toxin [Streptococcus sp. HSISS2]** | **Streptococcus sp. HSISS2** | **229** | **229** | **0.99** | **4E-75** | **0.9187** | **123** | **EQC72467.1** |
| KJE23_02395 | hypothetical protein | | | | | | | | | |
| KJE23_02400 | hypothetical protein | | | | | | | | | |
| KJE23_02405 | AAA family ATPase | | | | | | | | | |
| KJE23_02445 | hypothetical protein | | | | | | | | | |
| KJE23_02920 | CPBP family intramembrane metalloprotease | | | | | | | | | |
| KJE23_02925 | helix-turn-helix domain-containing protein | | | | | | | | | |
| KJE23_03255 | bacteriophage abortive infection AbiH family protein | | | | | | | | | |
| KJE23_03265 | hypothetical protein | | | | | | | | | |

| | |
|---|---|
| KJE23_03270 | hypothetical protein |
| KJE23_04425 | CRISPR-associated helicase/endonuclease Cas3 |
| KJE23_04430 | type I-E CRISPR-associated protein Cse1/CasA |
| KJE23_04435 | type I-E CRISPR-associated protein Cse2/CasB |
| KJE23_04440 | type I-E CRISPR-associated protein Cas7/Cse4/CasC |
| KJE23_04445 | type I-E CRISPR-associated protein Cas5/CasD |
| KJE23_04450 | type I-E CRISPR-associated protein Cas6/Cse3/CasE |
| KJE23_04455 | type I-E CRISPR-associated endonuclease Cas1 |
| KJE23_04460 | type I-E CRISPR-associated endoribonuclease Cas2 |
| KJE23_04465 | hypothetical protein |
| KJE23_04795 | VWA domain-containing protein |
| KJE23_05095 | hypothetical protein |
| KJE23_05100 | ABC transporter |
| KJE23_05280 | sulfurtransferase TusA family protein |
| KJE23_05285 | YeeE/YedE family protein |
| KJE23_06390 | ABC transporter ATP-binding protein/permease |
| KJE23_06395 | helix-turn-helix domain-containing protein |
| KJE23_06850 | MBL fold metallo-hydrolase |
| KJE23_06970 | helix-turn-helix domain-containing protein |
| KJE23_06975 | hypothetical protein |
| KJE23_07275 | hypothetical protein |
| KJE23_07280 | DUF2142 domain-containing protein |
| KJE23_07285 | glycosyltransferase |
| KJE23_07485 | AAA family ATPase |
| KJE23_08085 | hypothetical protein |
| KJE23_08400 | class IIb bacteriocin, lactobin A/cerein 7B family |

| | | | | | | | | | | |
|---|---|---|---|---|---|---|---|---|---|---|
| KJE23_09075 | low temperature requirement protein A | | | | | | | | | |
| KJE23_09260 | hypothetical protein | | | | | | | | | |
| KJE23_09615 | hypothetical protein | | | | | | | | | |
| KJE23_09620 | arsenical efflux pump membrane protein ArsB | | | | | | | | | |
| KJE23_09625 | exonuclease | | | | | | | | | |
| KJE23_09630 | hypothetical protein | | | | | | | | | |
| KJE23_09635 | molybdopterin-dependent oxidoreductase | | | | | | | | | |
| **KJE23_09650** | **FliM/FliN family flagellar motor switch protein** | **type III secretion system cytoplasmic ring protein SctQ [Lysobacter enzymogenes]** | **Lysobacter enzymogenes** | **159** | **159** | **0.33** | **3E-43** | **1** | **364** | **WP_096382596.1** |
| KJE23_09655 | hypothetical protein | | | | | | | | | |
| KJE23_09660 | DUF2292 domain-containing protein | | | | | | | | | |
| KJE23_09670 | protein-export chaperone SecB | | | | | | | | | |
| KJE23_09675 | hypothetical protein | | | | | | | | | |
| KJE23_09680 | host cell division inhibitory peptide Kil | | | | | | | | | |
| KJE23_09685 | hypothetical protein | | | | | | | | | |
| KJE23_09690 | hypothetical protein | | | | | | | | | |
| KJE23_09695 | hypothetical protein | | | | | | | | | |
| **KJE23_09700** | **hypothetical protein** | **O-antigen ligase family protein [Lysobacter sp. K5869]** | **Lysobacter sp. K5869** | **76.6** | **76.6** | **0.28** | **1E-12** | **0.6818** | **462** | **WP_215837858.1** |
| KJE23_09705 | hypothetical protein | | | | | | | | | |
| KJE23_09710 | hypothetical protein | | | | | | | | | |
| KJE23_09715 | DUF533 domain-containing protein | | | | | | | | | |
| KJE23_09725 | hypothetical protein | | | | | | | | | |
| **KJE23_09735** | **sigma 54-interacting transcriptional regulator** | **type 4 pilus PilR [Lysobacter enzymogenes]** | **Lysobacter enzymogenes** | **166** | **166** | **0.32** | **2E-44** | **0.988** | **471** | **QEU57250.1** |
| KJE23_09745 | hypothetical protein | | | | | | | | | |
| KJE23_09750 | hypothetical protein | | | | | | | | | |

| | |
|---|---|
| KJE23_09755 | S8 family serine peptidase |
| KJE23_09765 | outer membrane beta-barrel protein Lom |
| KJE23_09770 | transglycosylase SLT domain-containing protein |
| KJE23_09775 | hypothetical protein |
| KJE23_09780 | hypothetical protein |
| KJE23_09790 | phage tail length tape measure family protein |
| KJE23_09795 | S49 family peptidase |

Bold listings have annotations that could be considered virulent factors

A.

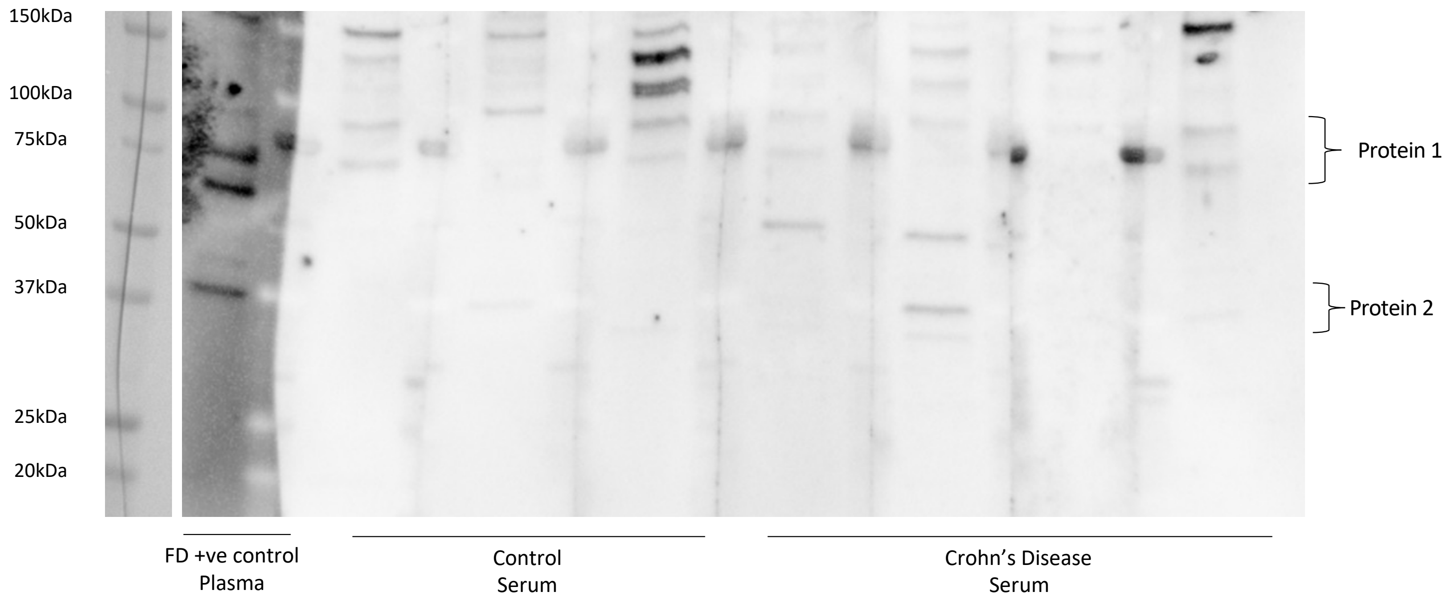
150kDa
100kDa
75kDa
50kDa
37kDa
25kDa
20kDa
Protein 1
Protein 2
FD +ve control
Plasma
Control
Serum
Crohn's Disease
Serum

B.

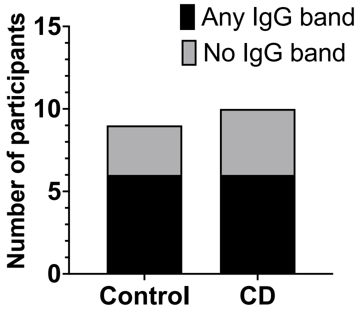
Any IgG band
No IgG band
Number of participants
15
10
5
0
Control
CD

C.

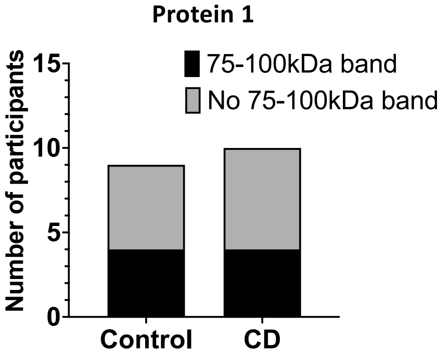
Protein 1
75-100kDa band
No 75-100kDa band
Number of participants
15
10
5
0
Control
CD

D.

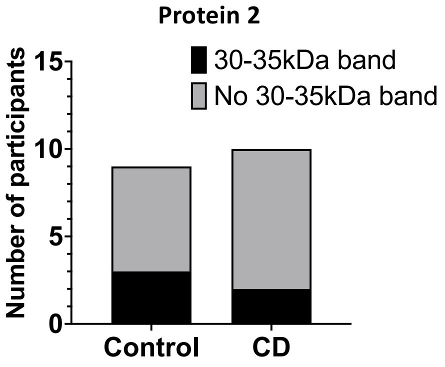
Protein 2
30-35kDa band
No 30-35kDa band
Number of participants
15
10
5
0
Control
CD

**Supplementary Figure 1: Screening of Streptococcus salivarius AGIRA0003 total protein lysates for IgG interactions with serum from Crohn's disease patients and controls.**

Total protein extracted from bacterial lysates was electrophoresed and immunoblotted with patient plasma as the probing antibody to determine if (A) the interaction between the duodenal microbiota and patient plasma was also observed in Crohn's disease (CD), as an example of an organic gastrointestinal disease. (B) The presence of such an interaction with *S. salivarius* AGIRA0003 at either molecular weight, (C) ~75-100kDa (Protein 1) or (D) ~30-35kDa (Protein 2) were tested for potential associations with Crohn's disease, n=9 controls, n=10 CD. Statistical analysis: Chi-Squared test.

Supplementary Figure 2

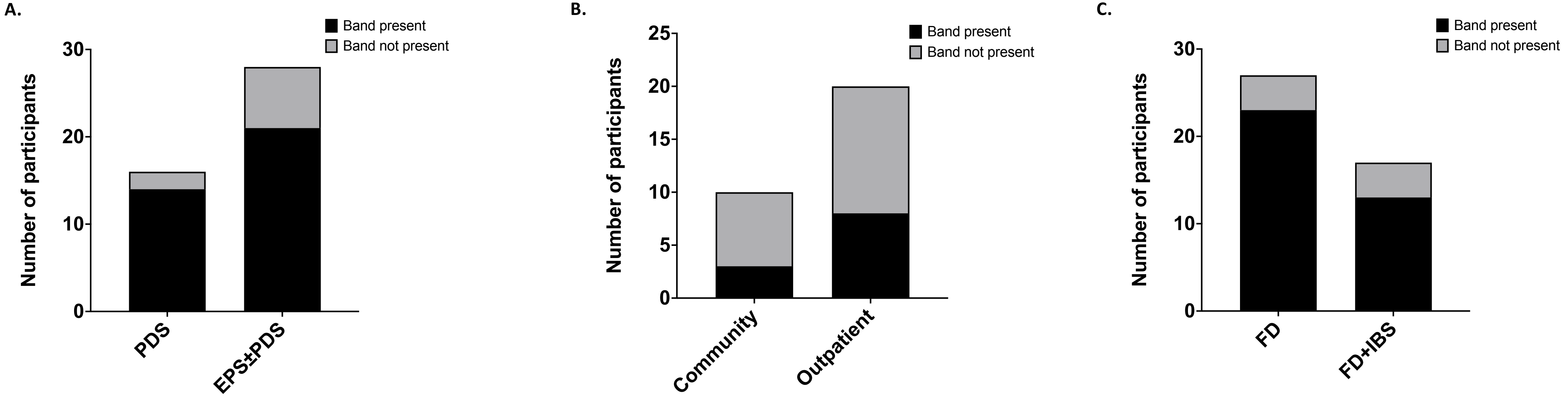

**Supplementary Figure 2: The IgG response against *Streptococcus salivarius* AGIRA0003 by FD subtype and control type.**

The presence of an interaction with *S. salivarius* AGIRA0003 was assessed by Chi-square testing between (A) FD patients with the post-prandial distress subtype compared to those with epigastric pain with or without post-prandial distress symptoms; as well as between (B) controls recruited from the community as healthy subjects compared to those recruited from the outpatient setting and between (C) patients with FD only versus those with FD and IBS overlap. n=16 PDS, n=28 EPS±PDS, n=10 community controls, n=20 outpatient controls, n=27 FD, n=17 FD+IBS.

Supplementary Figure 3

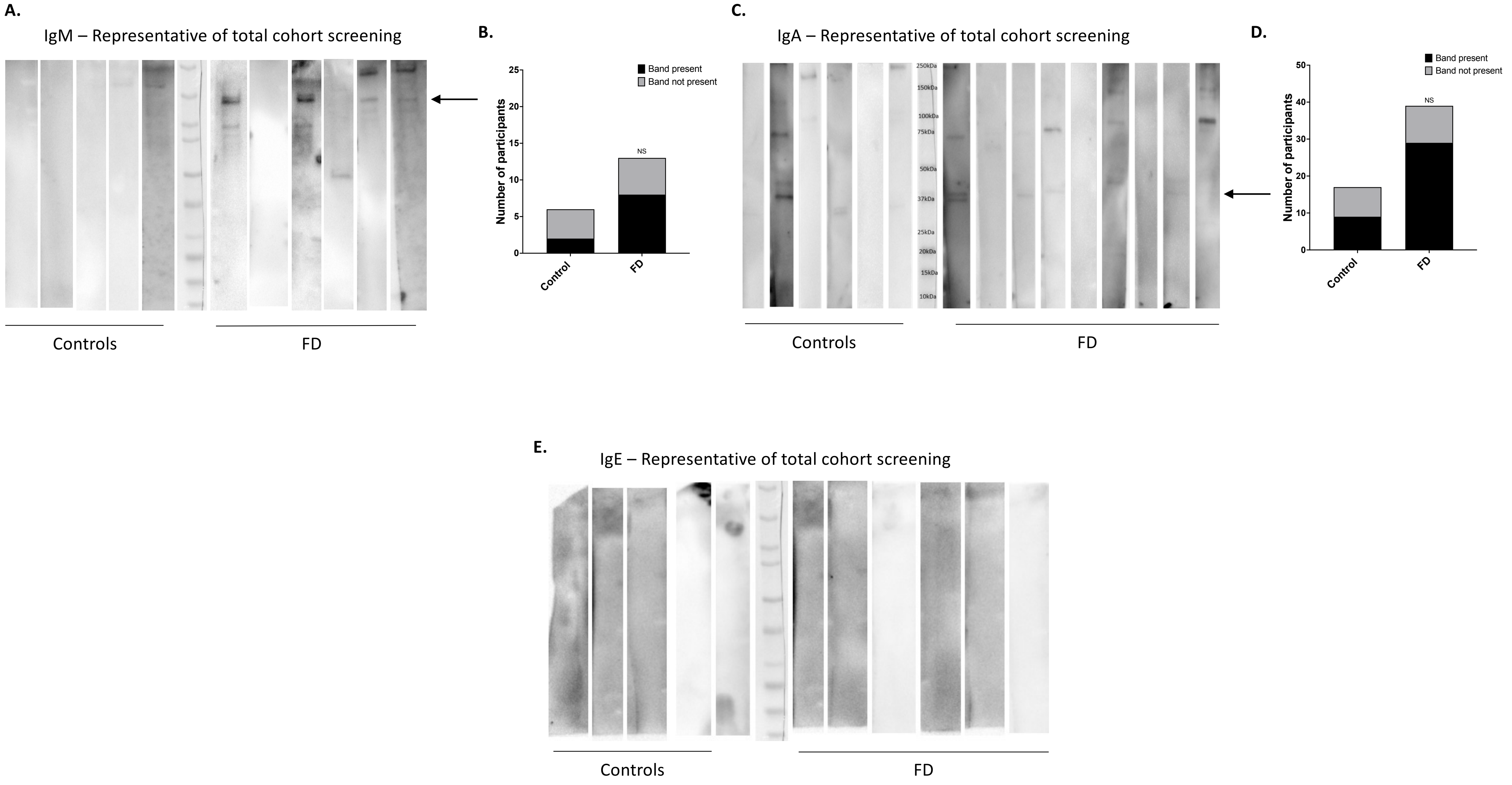

**Supplementary Figure 3: Screening of *Streptococcus salivarius* AGIRA0003 total protein lysates for IgM, IgA and IgE antibodies in plasma of controls and FD patients.** Patients were screened for (A) IgM and (B) the proportions of IgM seroreactive FD patients and controls were analysed. (C) Patient plasma was screened for IgA antibodies against proteins from *Streptococcus salivarius* AGIRA0003 and (D) tested for associations with FD or controls. (E) No IgE response was detected in any of the controls or patients screened. n=6 controls and n=13 FD for (B), n=17 controls and n=39 FD for (D) and n=8 controls and n=21 FD for (E). Statistical analysis: Chi-square test.

Supplementary Figure 4

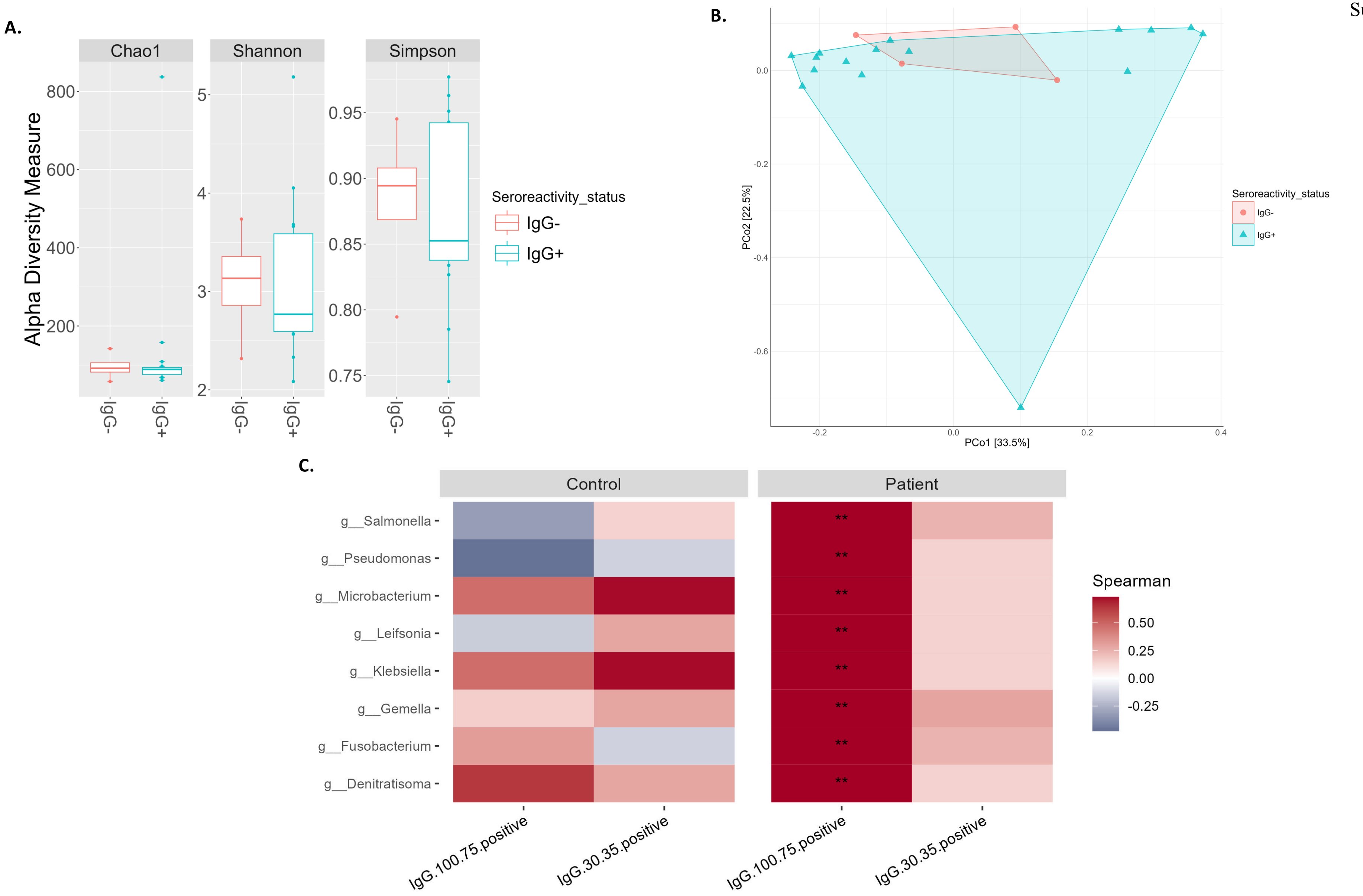

**Supplementary Figure 4: The duodenal microbiota of patients with and without IgG antibodies against *Streptococcus salivarius* AGIRA0003.**

(A) Alpha diversities represented by Chao1, Shannon and Simpson indices for samples $IgG^+$ and $IgG^-$ to S. salivarius AGIRA0003, irrespective of control or FD status. (B) Principal coordinates analysis plot of Bray-Curtis beta diversity of the same samples. (C) Heatmap of Spearman's correlation of bacteria genus correlated with FD patient *S. salivarius* AGIRA0003 seroreactivity status compared to controls. Red indicates positive correlation, blue equals negative correlation.

Supplementary Figure 5

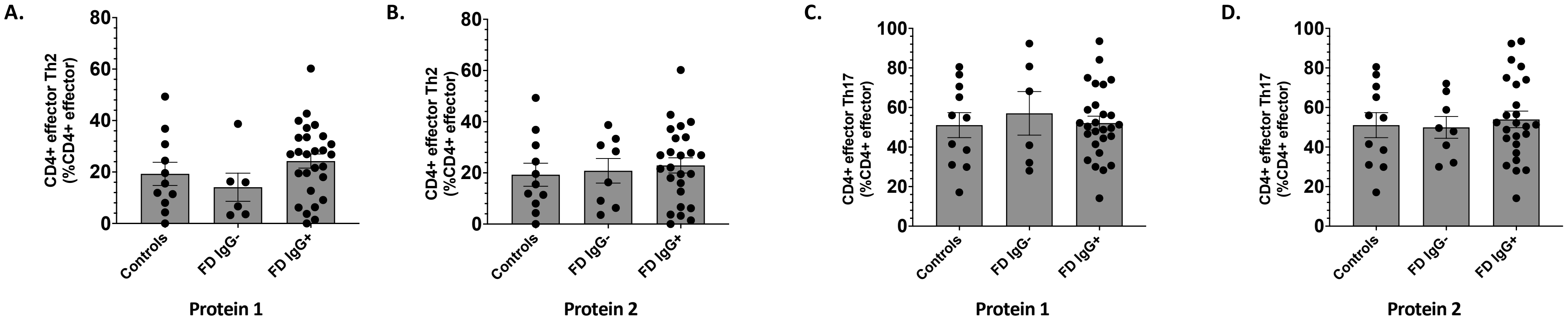

**Supplementary Figure 5: IgG seroreactive status and duodenal effector T cells in FD patients.**

Duodenal lymphocytes analysed from a subset of patients and controls by flow cytometry in a previous study were investigated between FD patients with and without IgG seroreactivity and controls. The proportion of effector T helper (Th)-2 like cells was compared against IgG status for both (A) Protein 1 and (B) Protein 2. Effector Th17-like cells were also compared against IgG status for both (C) Protein 1 and (D) Protein 2. n=11 controls, n=6 $IgG^-$ FD for Protein 1, n=27 $IgG^+$ FD for Protein 1, n=8 $IgG^-$ FD for Protein 2, n=25 $IgG^+$ FD for Protein 2. Data presented as mean±SEM. Statistical analysis: (A, C) non-parametric one-way ANOVA, (B, D) parametric one-way ANOVA.

Supplementary Figure 6

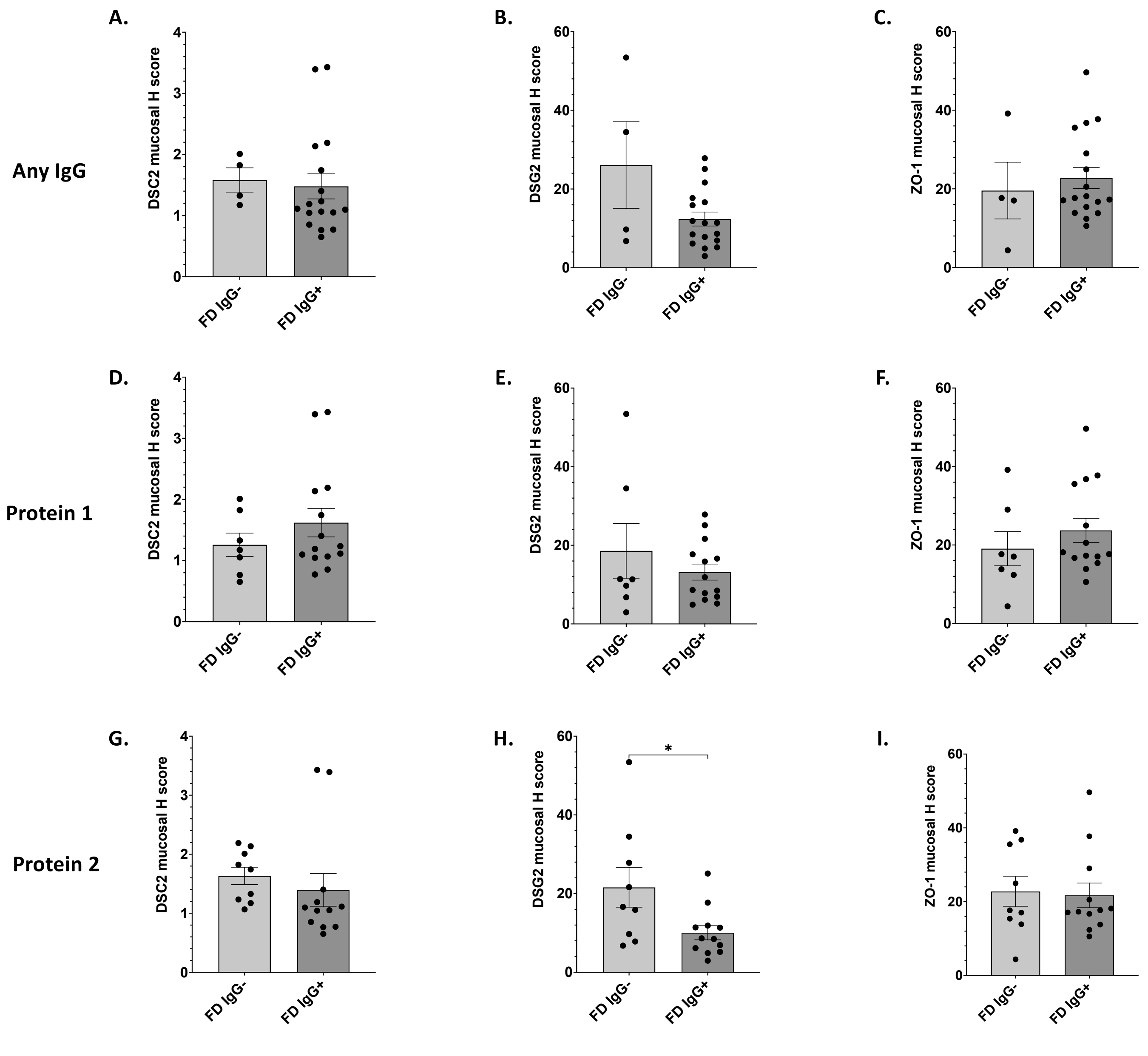

**Supplementary Figure 6: Tight junction associated proteins, DSC2, DSG, and ZO-1, in duodenal biopsies from IgG$^{+}$ FD patients compared to IgG$^{-}$ FD patients.**

Formalin fixed, paraffin embedded duodenal biopsies immunohistochemically stained with DSC2, DSG2 and ZO-1 were analysed for IgG$^{+}$ FD patients compared to IgG$^{-}$ FD patients. H-scores were compared for (A) DSC2, (B) DSG2 and (C) ZO-1 in patients positive for either Protein 1 and/or Protein 2. (D) DSC2, (E) DSG2 and (F) ZO-1 levels were also compared between patients IgG+ and IgG- for Protein 1, as well as Protein 2 individually ((G) DSC2, (H) DSG2 and (I) ZO-1). (A-C) n=4 IgG$^{-}$, n=17 IgG$^{+}$ FD. (D-F) n=7 IgG$^{-}$, n =14 IgG$^{+}$ FD. (G-I) n=9 IgG$^{-}$, n=12 IgG$^{+}$ FD. Data presented as mean±SEM. Statistical analysis: non-parametric one-way ANOVA. *$p$<0.05.
